# Molecular and clinical signatures in Acute Kidney Injury define distinct subphenotypes that associate with death, kidney, and cardiovascular events

**DOI:** 10.1101/2021.12.14.21267738

**Authors:** George Vasquez-Rios, Wonsuk Oh, Samuel Lee, Pavan Bhatraju, Sherry G. Mansour, Dennis G. Moledina, Heather Thiessen-Philbrook, Eddie Siew, Amit X. Garg, Vernon M. Chinchilli, James S. Kaufman, Chi-yuan Hsu, Kathleen D. Liu, Paul L. Kimmel, Alan S. Go, Mark M. Wurfel, Jonathan Himmelfarb, Chirag R. Parikh, Steven G. Coca, Girish N. Nadkarni

## Abstract

**Introduction:** AKI is a heterogeneous syndrome defined via serum creatinine and urine output criteria. However, these markers are insufficient to capture the biological complexity of AKI and not necessarily inform on future risk of kidney and clinical events.

**Methods:** Data from ASSESS-AKI was obtained and analyzed to uncover different clinical and biological signatures within AKI. We utilized a set of unsupervised machine learning algorithms incorporating a comprehensive panel of systemic and organ-specific biomarkers of inflammation, injury, and repair/health integrated into electronic data. Furthermore, the association of these novel biomarker-enriched subphenotypes with kidney and cardiovascular events and death was determined. Clinical and biomarker concentration differences among subphenotypes were evaluated via classic statistics. Kaplan-Meier and cumulative incidence curves were obtained to evaluate longitudinal outcomes.

**Results:** Among 1538 patients from ASSESS-AKI, we included 748 AKI patients in the analysis. The median follow-up time was 4.8 years. We discovered 4 subphenotypes via unsupervised learning. Patients with AKI subphenotype 1 (‘injury’ cluster) were older (mean age ± SD): 71.2 ± 9.4 (p<0.001), with high ICU admission rates (93.9%, p<0.001) and highly prevalent cardiovascular disease (71.8%, p<0.001). They were characterized by the highest levels of KIM-1, troponin T, and ST2 compared to other clusters (P<0.001). AKI subphenotype 2 (‘benign’ cluster) is comprised of relatively young individuals with the lowest prevalence of comorbidities and highest levels of systemic anti-inflammatory makers (IL-13). AKI Subphenotype 3 (‘chronic inflammation and low injury’) comprised patients with markedly high pro-BNP, TNFR1, and TNFR2 concentrations while presenting low concentrations of KIM-1 and NGAL. Patients with AKI subphenotype 4 (‘inflammation-injury’) were predominantly critically ill individuals with the highest prevalence of sepsis and stage 3 AKI. They had the highest systemic (IL-1B, CRP, IL-8) and kidney inflammatory biomarker activity (YKL-40, MCP-1) as well as high kidney injury levels (NGAL, KIM-1). AKI subphenotype 3 and 4 were independently associated with a higher risk of death compared to subphenotype 2. Moreover, subphenotype 3 was independently associated with CKD outcomes and CVD events.

**Conclusion:** We discovered four clinically meaningful AKI subphenotypes with statistical differences in biomarker composites that associate with longitudinal risks of adverse clinical events. Our approach is a novel look at the potential mechanisms underlying AKI and the putative role of biomarkers investigation.

## Introduction

Acute kidney injury (AKI) is a heterogeneous syndrome that may occur in the context of diverse clinical scenarios including infection, acute organ failure, or hemodynamic changes, among others.^1, 2^ Despite the broad definition, clinicians intuitively have classified AKI into ‘pre-renal’, ‘renal’ (or intrinsic), and ‘post-renal’ causes.^1, 3^ However, given the complexity of such disease processes, multiple insults may co-exist and induce different biological disturbances leading to AKI and adverse outcomes. Ascertainment of pathological pathways, duration of disease, and prediction of clinical events is difficult via traditional measures such as serum creatinine fluctuations, urine studies, and urine output changes.^4^ Furthermore, kidney function following AKI in the short to medium term can display several non-linear trajectories over time (including ongoing recovery) that may not associate with meaningful laboratory abnormalities, nor the etiology of AKI adjudicated during the acute hospitalization.^1, 4–6^ This represents a major limitation to providing individualized care during the acute setting and in the post-AKI stage.

Biomarkers reflective of key biological pathways such as kidney injury, inflammation, and health have emerged over the past 10 years showing their potential to anticipate the diagnosis of acute kidney injury (AKI) and forecast adverse outcomes in this population including disease duration, dialysis needs, and mortality.^7, 8^ Furthermore, among CKD patients, some of these biomarkers have been demonstrated to strongly predict the advent of end-stage kidney disease (ESKD) in patients with and without diabetes mellitus.^9–15^ However, their ability to outperform to less expensive and routinely collected laboratory tests such as creatinine and urine albumin-to-creatinine ratio (uACR) has been questioned.^16^ While efforts have been made to integrate individual biomarkers (or panels) into clinical prediction models, standard regression models may be limited to evaluate markedly heterogeneous and high-dimensional data, thereby compromising the researcher’s ability to find clinical significance from these. Importantly, the importance of incorporating ‘big data’ to enrich our perspective on AKI has been recently acknowledged 15^th^ Acute Dialysis Quality Initiative (ADQI).^17^

Unsupervised learning plays an important role in data summarization and preliminary structure identification of complex and highly variable data.^18, 19^ Considering the complex nature of AKI, contributing clinical factors, and implicated mechanisms, unsupervised clustering has the potential to serve as an informative analytical tool to identify AKI subphenotypes from high-dimensional data.^17, 20, 21^ Previous studies suggested that AKI subphenotypes based upon tumor necrosis factor receptors (TNFR) and angiopoietin-2/angiopoietin-1 ratio could serve to identify patients’ responsiveness to vasopressors and short-term adverse outcomes in a critically ill population via latent class analyses.^22^ Furthermore, our group demonstrated that unsupervised learning (deep learning) integrating routine laboratory parameters and electronic health record (EHR) data could lead to subphenotypes associated with short-term kidney outcomes in a sepsis-AKI population.^23^ However, whether a comprehensive panel of kidney and systemic biomarkers reflective of different axes of health and disease could serve to identify agnostic clinical and biological signatures in a broader population of AKI and uncover potential associations with longitudinal clinical events if unknown.

The Assessment, Serial Evaluation, and Subsequent Sequelae in Acute Kidney Injury (ASSESS-AKI was a prospective cohort of AKI survivors whose clinical and biomarker data was rigorously collected on admission and in a protocolized fashion after discharge.^24^ While data on an individual or small set of post-operative biomarkers and their associations with AKI and CKD are extensive in the literature,^25–27^ adequate integration of multiple systemic and kidney-related biomarkers of inflammation, injury, and fibrosis along with clinical data to investigate distinct AKI profiles with clinical significance and potential associations with long-term outcomes is lacking. We sought to discover AKI subphenotypes through a series of unsupervised machine learning algorithms (hierarchical agglomerative clusters, and K-means clustering) and validation models (consensus clustering). We also aimed to identify whether these novel clusters could associate with longitudinal kidney, cardiovascular events, and death.

## Methods

### Cohort description and covariates

The ASSESS-AKI study was a multicenter, prospective, matched parallel cohort of 1538 hospitalized adult participants who developed AKI and matched individuals who did not have AKI and who survived to complete an in-person visit at 3 months after discharge.^24^ Patients aged 18 and over were enrolled from the medical and surgical floors and intensive care units (ICUs) in 4 US-based medical centers with matching criteria at each institution for baseline covariates, estimated glomerular filtration rate (eGFR), urine albumin-to-creatinine (uACR) levels and AKI stage. Patients had pre-admission serum creatinine measurements obtained within 1 year of the index hospitalization as outpatients. The etiology of AKI was not adjudicated during the hospitalization. Exclusion criteria for ASSESS-AKI were broad and included the presence of acute glomerulonephritis, hepatorenal syndrome, multiple myeloma, malignancy, urinary obstruction, severe heart failure, kidney replacement therapy (dialysis, transplant) prior to hospitalization, pregnancy, or predicted survival ≤ 12 months. For the present study, the focus of our analyses were those patients who had AKI as defined by the ASSESS-AKI protocol during the hospitalization who survived 3-months after discharge (N=769).

AKI was defined as a relative increase of ≥50% or absolute increase of ≥ inpatient serum creatinine concentration above the baseline creatinine. AKI stage was classified following the Acute Kidney Injury Network (AKIN) guidelines. CKD status and eGFR calculation were obtained using the Chronic Kidney Disease Epidemiology Collaboration (CKD-EPI) equation. Participants had study visits at 3 and 12 months after discharge, and annually thereafter (interim phone contacts every 6 months) for assessment of biomarker data and adjudication of clinical events. Variables including demographic characteristics and comorbidities were obtained from medical records. Serum creatinine was measured using an isotope dilution mass spectrometry-traceable enzymatic assay (Roche Di-agnostics, Indianapolis, IN), and a random spot urine protein-to-creatinine ratio using a turbidimetric method (Roche). Biomarker quartiles were defined within the AKI cohort via different diagnostic assays.

### Biomarker measurements

Plasma and urine samples were collected within 96 hours of the diagnosis of AKI. Systemic biomarkers of inflammation included: IFN-γ, IL-4, IL-13, TNF-α, IL-1β, IL-2, IL-6, IL-8, IL-10, and IL-12. Kidney biomarkers of inflammation were: TNFR1, TNFR2, YLK-40, MCP-1, and IL-18, whereas biomarkers of kidney injury included: KIM-1, NGAL, uACR. UMOD was evaluated to inform on tubule health. Biomarkers of cardiac congestion, injury, and remodeling/fibrosis, respectively, included: pro-BNP, Troponin T, ST2 and Gal-3. Blood samples were collected in ethylenediamine tetra-acetic acid tubes and centrifuged to separate plasma. Sample underwent a single controlled thaw, were centrifuged at 5000xg for 10 minutes at 4*C, separated into 1 mL aliquots, and immediately stored at -80°C until biomarkers were measured. Urine biomarkers were obtained in a protocolized fashion and measured via multiplex assay or by using Meso Scale Diagnostics when appropriate. Details on biomarker collection and processing have been described previously.^26^

### Outcome definitions

We evaluated three main outcomes: Composite kidney outcome (CKD incidence or progression), cardiovascular events, and death. Among patients without underlying CKD, CKD incidence was defined as >25% reduction in the eGFR from baseline; or reaching CKD 3 or worse during follow-up. Among patients with underlying CKD (eGFR <60 ml/min/1.73m^2^ during hospitalization), CKD progression was ascertained as >50% reduction in the baseline eGFR; or progression to CKD 5 or development of ESKD on follow-up (hemodialysis or peritoneal dialysis requirement at least once/week for >12 weeks, receiving a kidney transplant and/or death while on dialysis). Cardiovascular events were a composite of myocardial infarction, heart failure, cerebrovascular accidents, peripheral artery disease or coronary or cardiovascular intervention. Death was ascertained from proxy reports, medical records, and death certificate data.

### Data preprocessing and subphenotype discovery

Patients’ demographics, comorbidities and a comprehensive panel of systemic and organ-specific biomarkers were included during data preparation. First, for numerical variables, outliers were adjusted using 95% winsorization. In our study, observations greater than 97.5% were set to 97.5%, and observations smaller than 2.5% were set to 2.5%. Second, variables showing left-skewed distribution were log-transformed and examined whether the mode leans to the median than the first quartile. Third, variables were normalized using a robust scaler,^28^ to attenuate the influence of observations with extreme values. Finally, multivariate imputation by chained equations (MICE)^29, 30^ was used with 5 multiple imputations and 50 iterations. Creating multiple imputations, as opposed to single imputations accounts for the statistical uncertainty in the imputations. In addition, the chained equations approach is very flexible and can handle a wide array of variables.

After numerical variables were cleaned and normalized, both categorical and numerical variables were transformed into lower-dimensional space using factor analysis of mixed data (FAMD) (ref). The use of a total of 53 variables can hurt clustering through two perspectives; spareness of variables, which is known as a curse of dimensionality, and multicollinearity. While several methods exist for dimensionality reduction to address these challenges, such as matrix factorization^31, 32^ and deep autoencoder,^33, 34^ FAMD is a recently developed principal component analysis – family method that simultaneously explores multivariate dependencies between both categorical and numerical variables. 14 out of 53 principal components (eigenvalues >1) were selected based on the Kaiser-Guttman criterion.

We conducted hierarchical agglomerative clustering in which each observation starts in its own cluster and pairs of clusters are merged as one moves up the hierarchy. We used ward linkage with Euclidean distance applied to 14 principal components. The number of subphenotypes was determined through the visual evaluation of the dendogram as well as results from 26 indexes available from the NbClust package in R.^35^ Furthermore, the robustness of our clustering algorithms was evaluated through alternative techniques including K-means and consensus clustering. After obtention of clusters, we described the clinical and biomarker distribution per cluster group. Final Shapley additive explanations (SHAP) diagrams were obtained to assess the importance of intervening features on each subphenotype by comparing the XGBoost models with and without the presence of such specific feature.

### Statistical analysis

Descriptive statistics for continuous variables were reported as mean (standard deviation) or median (interquartile range) accordingly for each of the subphenotypes. Categorical variables were presented as frequencies and percentages. Clinical, demographic and biomarker concentration differences among subphenotypes were evaluated via the analysis of variance (ANOVA) or Chi-square test when appropriate. Cox proportional hazard models were used compare time to censored kidney events (CKD incidence or progression), CVD events and death with death up to 6 years of follow-up. These models were fully adjusted for age, gender, diabetes mellitus (yes/no), BMI, baseline CKD (yes/no), and baseline UACR. Two-tailed P values of less than 0.05 were considered to indicate statistical significance. We performed all statistical analyses using R software version 4.0.3 (R Foundation for Statistical Computing, Vienna, Austria) and Python 3.9.3.

## RESULTS

### Clinical features of AKI patients

Among 1538 participants included in ASSESS-AKI, 748 AKI individuals had full biomarker and data of interest and were included in the analysis. The median follow-up time for clinical events was 4.8 years. Participants were predominantly men (68%), from Caucasian ethnicity (79%) with a mean (± SD) of 64 (13). The prevalence of baseline for key covariates such as CKD (eGFR<60 ml/min/1.73 m^2^), cardiovascular disease, congestive heart failure (CHF), hypertension and diabetes mellitus were: 39.7%, 48%, 26.3%, 78.6% and 50%, respectively. The overall mean (± SD) baseline eGFR was 66.8 (24.9) ml/min per 1.73m^2^ and the overall mean (± SD) uACR was 2.2 mg/g in this population. A large proportion of the cases of AKI were stage 1 (72.6%), while AKI stage 2 (15%) and 3 (12.4%) were less frequent. Full baseline and AKI clinical characteristics are reflected in **Table 1**.

**Table 1.** Clinical characteristics among the four novel AKI subphenotypes

|  | **1 (N=181)** | **2 (N=250)** | **3 (N=159)** | **4 (N=158)** | **p value** |
| --- | --- | --- | --- | --- | --- |
| **Age** |  |  |  |  | < 0.001 |
| **Mean (SD)** | 71.2 (9.4) | 60.2 (12.8) | 66.8 (11.6) | 57.4 (12.4) |  |
| **Range** | 32.7 - 87.3 | 18.8 - 82.7 | 35 - 88.9 | 22.2 - 87.8 |  |
| **Gender** |  |  |  |  | < 0.001 |
| **M** | 148 (81.8%) | 162 (64.8%) | 104 (65.4%) | 94 (59.5%) |  |
| **F** | 33 (18.2%) | 88 (35.2%) | 55 (34.6%) | 64 (40.5%) |  |
| **Race** |  |  |  |  | < 0.001 |
| **W** | 174 (96.1%) | 180 (72.0%) | 112 (70.4%) | 128 (81.0%) |  |
| **B** | 5 (2.8%) | 57 (22.8%) | 38 (23.9%) | 11 (7.0%) |  |
| **O** | 2 (1.1%) | 13 (5.2%) | 9 (5.7%) | 19 (12.0%) |  |
| **ICU adm** |  |  |  |  | < 0.001 |
| **N** | 11 (6.1%) | 100 (40%) | 64 (40.3%) | 43 (27.2%) |  |
| **Y** | 170 (93.9%) | 150 (60%) | 95 (59.7%) | 115 (72.8%) |  |
| **HTN** |  |  |  |  | < 0.001 |
| **N** | 42 (23.2%) | 51 (20.4%) | 19 (11.9%) | 48 (30.4%) |  |
| **Y** | 139 (76.8%) | 199 (79.6%) | 140 (88.1%) | 110 (69.6%) |  |
| **DM** |  |  |  |  | < 0.001 |
| **N** | 111 (61.3%) | 131 (52.4%) | 47 (29.6%) | 84 (53.2%) |  |
| **Y** | 70 (38.7%) | 119 (47.6%) | 112 (70.4%) | 74 (46.8%) |  |
| **CVD** |  |  |  |  | < 0.001 |
| **N** | 51 (28.2%) | 167 (66.8%) | 59 (37.1%) | 112 (70.9%) |  |
| **Y** | 130 (71.8%) | 83 (33.2%) | 100 (62.9%) | 46 (29.1%) |  |
| **CHF** |  |  |  |  | < 0.001 |
| **N** | 138 (76.2%) | 208 (83.2%) | 71 (44.7%) | 134 (84.8%) |  |
| **Y** | 43 (23.8%) | 42 (16.8%) | 88 (55.3%) | 24 (15.2%) |  |
| **Sepsis** |  |  |  |  | < 0.001 |
| **N** | 175 (96.7%) | 228 (91.2%) | 153 (96.2%) | 77 (48.7%) |  |
| **Y** | 6 (3.3%) | 22 (8.8%) | 6 (3.8%) | 81 (51.3%) |  |
| **CKD** |  |  |  |  | < 0.001 |
| **N** | 113 (62.4%) | 205 (82.0%) | 16 (10.1%) | 117 (74.1%) |  |
| **Y** | 68 (37.6%) | 45 (18.0%) | 143 (89.9%) | 41 (25.9%) |  |
| **Baseline Cr** |  |  |  |  | < 0.001 |
| **Mean (SD)** | 1.1 (0.3) | 1 (0.3) | 1.9 (0.6) | 1.1 (0.4) |  |
| **Range** | 0.5 - 1.9 | 0.5 - 2.1 | 0.7 - 2.7 | 0.5 - 2.7 |  |
| **Baseline eGFR** |  |  |  |  | < 0.001 |
| **Mean (SD)** | 68.1 (18.4) | 77.9 (19.6) | 38.9 (17) | 75.784 (24.7) |  |
| **Range** | 33.1 - 115.3 | 31.2 - 117.9 | 17.7 - 109.2 | 22.8 - 117.9 |  |

**Table 1. Clinical characteristics among the four novel AKI subphenotypes (continued)**
| **AKI stage** |  |  |  |  |  |
| --- | --- | --- | --- | --- | --- |
| **1** | 165 (91.2%) | 205 (82.0%) | 119 (74.8%) | 54 (34.2%) |  |
| **2** | 13 (7.2%) | 37 (14.8%) | 15 (9.4%) | 47 (29.7%) |  |
| **3** | 3 (1.7%) | 8 (3.2%) | 25 (15.7%) | 57 (36.1%) |  |
| **Peak Cr** |  |  |  |  | < 0.001 |
| **Mean (SD)** | 1.7 (0.5) | 1.7 (0.5) | 3.4 (1.5) | 3.2 (1.9) |  |
| **Range** | 0.9 - 3.8 | 0.9 - 4.7 | 1.2 - 7.4 | 0.9 - 7.4 |  |
| **Peak eGFR** |  |  |  |  | < 0.001 |
| **Mean (SD)** | 41.2 (13.2) | 44.1 (12.8) | 21.082 (10.1) | 26.108 (15.2) |  |
| **Range** | 15.2 - 67.1 | 12.7 - 67.1 | 5 - 63.1 | 5 - 67.1 |  |
| **uACR at AKI** |  |  |  |  | < 0.001 |
| **Mean (SD)** | 0.8 (1.3) | 0.6 (1.1) | 5 (7.7) | 3.5 (5.7) |  |
| **Range** | 0 - 10.8 | 0 - 7.2 | 0 - 24.9 | 0.1 - 24.9 |  |

### Unsupervised clustering analysis

Using the 53 biomarker and clinical characteristics, the hierarchical clustering algorithm identified four clusters that best represented the data patterns of our AKI population. A diagrammatic representation (dendogram) used to ascertain the arrangement of the clusters produced by the corresponding analyses revealed four clusters (**Figure 1**). The Uniform Manifold Approximation and Projection (UMAP) was used for dimensionality reduction and cluster visualization revealing adequate independence (**Figure 1**). Additionally, the robustness of these four clusters was established by comparing with another unsupervised algorithm (K-means clustering); visualized through the heatmap and UMAP in **Figures 2 and Supplemental Figure 1.**, confirming the presence of 4 clusters. The presence and stability of four clusters was also reproduced through consensus clustering **(Supplemental Figure 2**).

**Figure 1.**
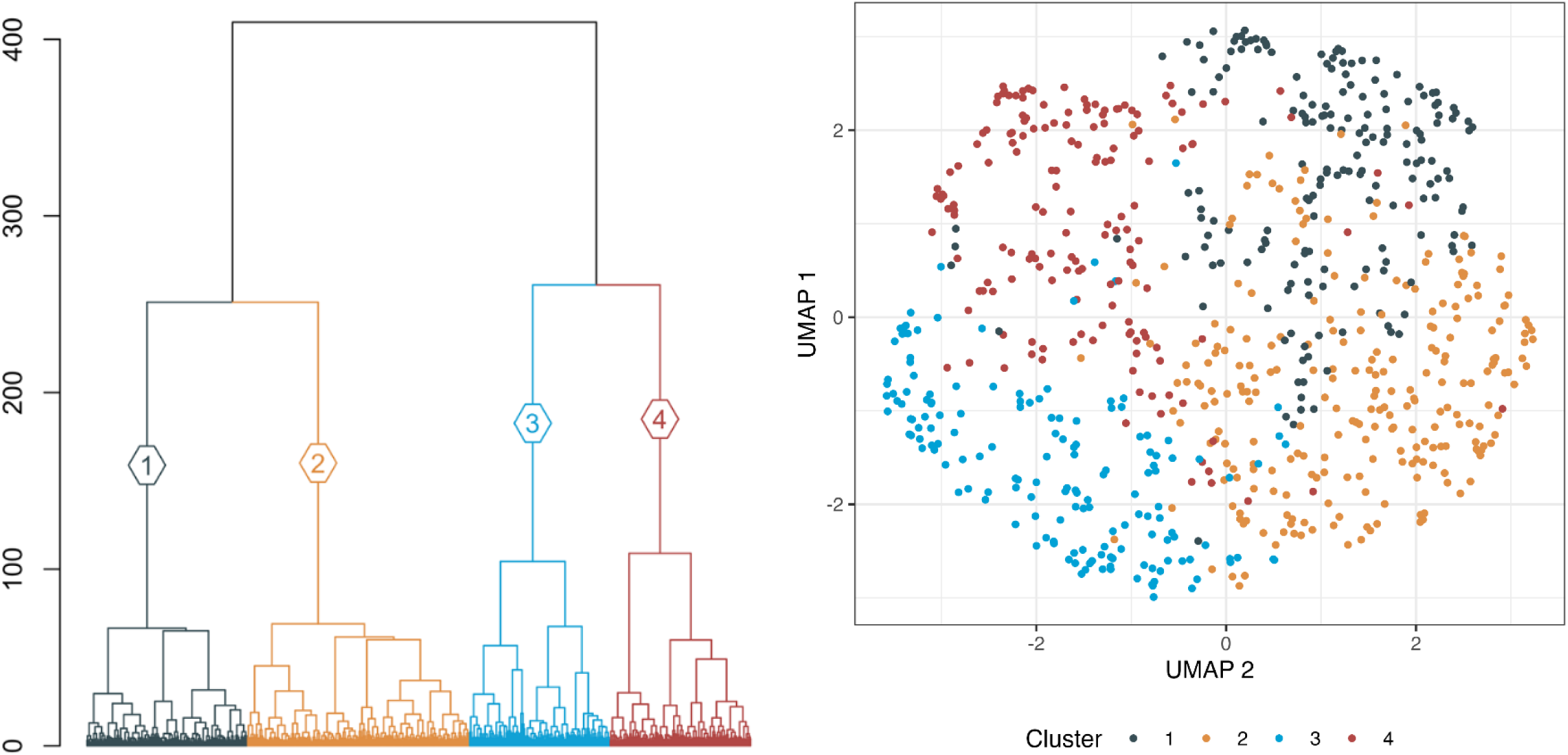
Hierarchical clustering, dendogram (left). UMAP showing density-based clustering and dimensionality reduction (right).

**Figure 2.**
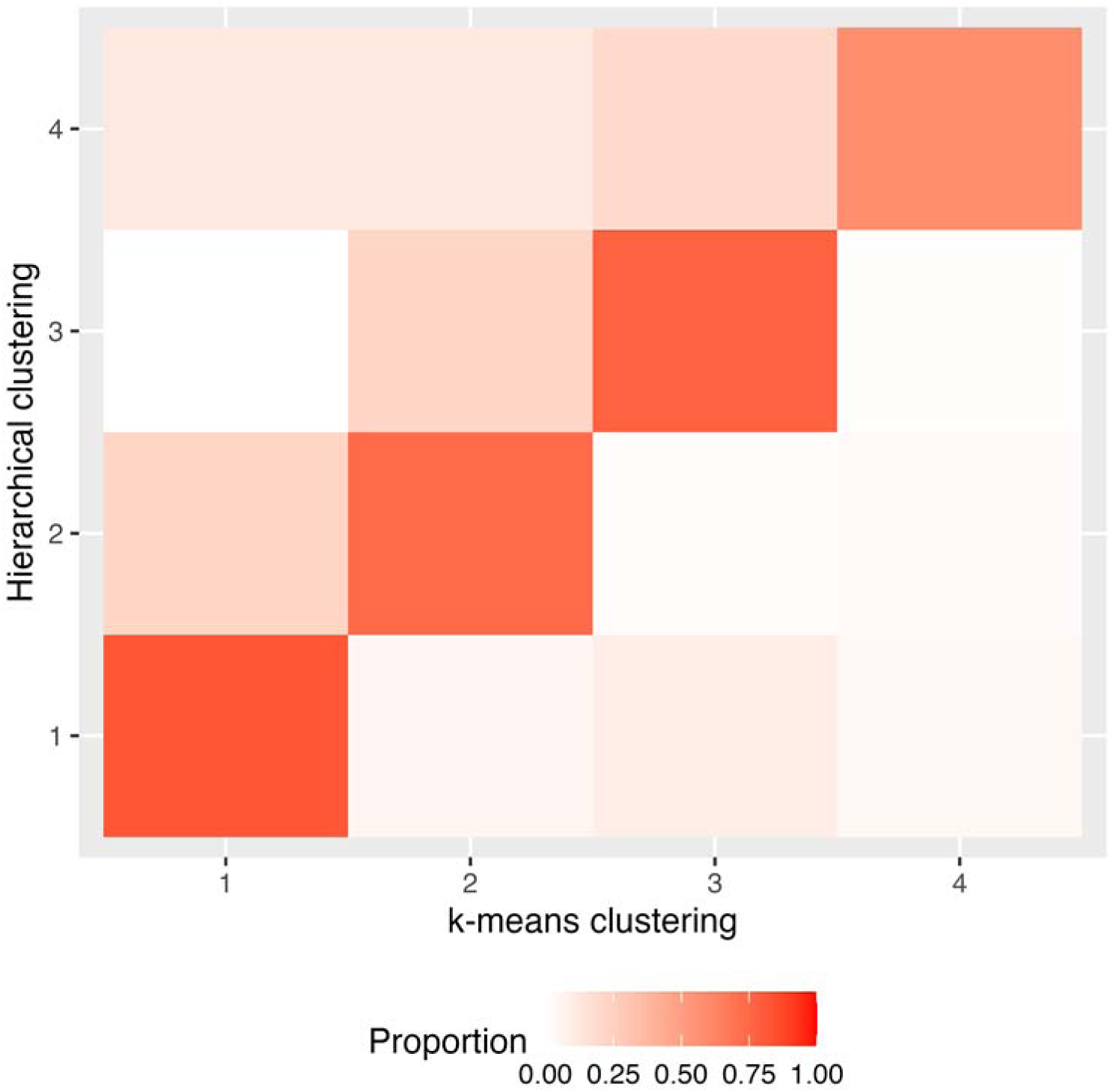
Heat map, robustness of 4 clusters.

### Clinical and biological signatures among AKI subphenotypes

The distributions of the 53 biomarker and clinical characteristics were statistically different between the four clusters (**Table 2**). Subphenotype 1 (N=181) was characterized by older individuals compared to subphenotype 2, 3 and 4, with age (mean ± SD): 71 ± 9 (p<0.001), with high ICU admission rates (94%, p<0.001), moderate rates of baseline CKD (38%, p<0.01) and the highest prevalence of baseline cardiovascular disease (72%, p<0.001) among all subphenotypes. Mean (SD) peak creatinine level at the time of diagnosis of AKI was 1.1 (0.3). Furthermore, individuals in subphenotype 1 had the highest rates of stage 1 AKI (91%). Subphenotype 2 (N=250) characterized by young individuals with the lowest prevalence of comorbid conditions such as CKD (18%, p<0.001), baseline CVD (33%, p<0.001), and CHF (16.8%, p<0.001). Mean (SD) peak creatinine level at the time of diagnosis of AKI was 1 (0.3) at the time of diagnosis and it was predominantly stage 1 (82%). Individuals in subphenotype 3 (N=159) had the highest prevalence of CKD (89.9%, p<0.001), diabetes mellitus (70.4%, p<0.001) and CHF (55.3%, p<0.001) compared to other subphenotypes. These patients presented the highest mean (SD) peak serum creatinine (1.9 ± 0.6) and albuminuria (5 ± 7.7) during the hospitalization. Also, the frequency of AKI stage 1, 2 and 3 were 74.8%, 9.4% and 15.7% respectively within this subphenotype. Patients in subphenotype 4 had markedly high ICU admission rates (73%, p<001) and the highest incidence of sepsis (51%, p<0.001) among all clusters. Furthermore, these individuals had comparatively lower rates of CKD (25.9%, p<0.001) and baseline cardiovascular disease (29.1%, p<0.001) than subphenotype 3. Notably, approximately 1 in every 3 patients within this subphenotype had stage 3 AKI, with mean (SD) peak serum creatinine of 3.2 (1.8) and albuminuria levels at 3.473 g/mg (5.7). The characteristics within each AKI subphenotype are described in **Table 2**.

**Table 2.** Biomarker characteristics within each AKI subphenotypes (systemic inflammatory) (cardiac injury, congestion, and repair/fibrosis) (kidney inflammation, injury and health)

| **Subphenotype** | **1 (N=181)** | **2 (N=250)** | **3 (N=159)** | **4 (N=158)** | **p value** |
| --- | --- | --- | --- | --- | --- |
| **INF-gamma** |  |  |  |  | < 0.001 |
| **Mean (SD)** | 2.9(6.9) | 12.7 (34.3) | 8.4 (19.5) | 39.6 (67.6) |  |
| **Range** | 0.3 - 71.3 | 0.3 - 225.6 | 0.3 - 225.6 | 0.3 - 225.6 |  |
| **IL1b** |  |  |  |  | < 0.001 |
| **Mean (SD)** | 0.3 (0.4) | 0.3 (0.3) | 0.3 (0.2) | 0.5 (0.5) |  |
| **Range** | 0.1 - 1.9 | 0.1 - 1.9 | 0.1 - 1.9 | 0.1 - 1.9 |  |
| **IL2** |  |  |  |  | < 0.001 |
| **Mean (SD)** | 0.5 (0.6) | 0.4 (0.5) | 0.5 (0.5) | 0.9 (1.1) |  |
| **Range** | 0.2 - 4.1 | 0.2 - 4.1 | 0.2 - 4.1 | 0.2 - 4.0 |  |
| **IL6** |  |  |  |  | < 0.001 |
| **Mean (SD)** | 50.8 (47.4) | 13.9 (24.9) | 13.1 (19.6) | 48.5 (63.3) |  |
| **Range** | 0.5 - 210.8 | 0.5 - 210.8 | 0.5 - 185.9 | 0.5 - 210.8 |  |
| **IL8** |  |  |  |  | < 0.001 |
| **Mean (SD)** | 15.6 (14.1) | 15.1 (17.1) | 15.1 (13.8) | 22.6 (22.3) |  |
| **Range** | 2.9 - 89.9 | 2.9 - 89.9 | 2.9 - 89.9 | 2.9 - 89.9 |  |
| **IL10** |  |  |  |  | < 0.001 |
| **Mean (SD)** | 1.9 (2.4) | 1.217 (1.7) | 0.9 (0.9) | 2.9 (3.4) |  |
| **Range** | 0.2 - 11.5 | 0.2 - 11.5 | 0.180 - 5.3 | 0.180 - 11.5 |  |
| **IL12** |  |  |  |  | < 0.001 |
| **Mean (SD)** | 0.3 (0.2) | 0.253 (0.3) | 0.267 (0.2) | 0.474 (0.5) |  |
| **Range** | 0.057 - 1.681 | 0.057 - 1.681 | 0.057 - 1.681 | 0.057 - 1.681 |  |
| **IL13** |  |  |  |  | < 0.001 |
| **Mean (SD)** | 0.5 (0.3) | 0.6 (0.4) | 0.6 (0.4) | 0.7 (0.5) |  |
| **Range** | 0.1 - 1.8 | 0.1 - 1.8 | 0.1 - 1.8 | 0.1 - 1.8 |  |
| **TNF-alpha** |  |  |  |  | < 0.001 |
| **Mean (SD)** | 4.403 (1.9) | 4.817 (2.4) | 7.227 (2.922) | 8.9 (4.7) |  |
| **Range** | 1.6 - 14.3 | 1.6 - 17.6 | 2.9 - 17.9 | 1.6 - 17.9 |  |

**Table 2. Biomarker characteristics within each AKI subphenotypes (cardiac injury, congestion, and repair/fibrosis) - continued**
| **Subphenotype** | **1 (N=181)** | **2 (N=250)** | **3 (N=159)** | **4 (N=158)** | **p value** |
| --- | --- | --- | --- | --- | --- |
| **Pro-BNP** |  |  |  |  | < 0.001 |
| **Mean (SD)** | 4185.8 (3520.6) | 1349.8(1827.9) | 5175.1 (4719.5) | 3265.1 (3459.1) |  |
| **Range** | 114 - 15693.7 | 32.8 - 11820 | 63 - 15693.6 | 32.8 - 15693.6 |  |
| **Troponin T** |  |  |  |  | < 0.001 |
| **Mean (SD)** | 564.1 (648.5) | 195.3 (473.9) | 352.9 (653.4) | 74.7 (188.9) |  |
| **Range** | 13.3 - 2741.8 | 13.300 - 2741.750 | 13.300 - 2741.750 | 13.3 - 1683 |  |
| **ST2** |  |  |  |  | < 0.001 |
| **Mean (SD)** | 354.7 (334.4) | 120.6 (162.9) | 144.6 (219.2) | 413.2 (476.9) |  |
| **Range** | 28.1 - 1513.3 | 17.3 - 1005.6 | 17.3 - 1513.3 | 17.3 - 1513.3 |  |
| **Gal3** |  |  |  |  | < 0.001 |
| **Mean (SD)** | 18.814 (8.1) | 15.874 (6.7) | 30.759 (11.8) | 22.621 (10.6) |  |
| **Range** | 6.9 - 52.9 | 6.8 - 42.7 | 6.8 - 52.9 | 6.8 - 52.9 |  |

**Table 2. Biomarker characteristics within each AKI subphenotypes (kidney inflammation, injury and health) - continued**
| **Subphenotype** | **1 (N=181)** | **2 (N=250)** | **3 (N=159)** | **4 (N=158)** | **p value** |
| --- | --- | --- | --- | --- | --- |
| **TNFR1** |  |  |  |  | < 0.001 |
| **Mean (SD)** | 5657.6 (2630.7) | 3863.6 (1759.7) | 9087.1 (4376.7) | 8377.4 (4400.3) |  |
| **Range** | 1960 - 19682.8 | 1566.8 - 12611 | 3393 - 19682.8 | 1566.8- 19682.8 |  |
| **TNFR2** |  |  |  |  | < 0.001 |
| **Mean (SD)** | 10697.9 (4487.9) | 9097.5 (5065.3) | 17534.2 (7852.5) | 19377.3 (10739.1) |  |
| **Range** | 3255.4 - 30631 | 3255.4 - 40119 | 4476 - 41167.5 | 3255.4 - 41167.5 |  |
| **Urine IL18** |  |  |  |  | < 0.001 |
| **Mean (SD)** | 85.4 (74.6) | 43.7 (55.8) | 46.485 (67.6) | 138.4 (120.8) |  |
| **Range** | 2.9 - 407.5 | 2.876 - 407.5 | 2.9 - 383.1 | 2.9 - 407.5 |  |
| **KIM-1** |  |  |  |  | < 0.001 |
| **Mean (SD)** | 9724.6 (8412.5) | 2655.8 (2889.7) | 2615.1 (3431.8) | 6159.9 (6249) |  |
| **Range** | 654 - 29077.2 | 70.4 - 20292 | 70.4 - 29077.2 | 236 - 29077.2 |  |
| **MCP-1** |  |  |  |  | < 0.001 |
| **Mean (SD)** | 1049.4 (1136.9) | 582.6 (835.5) | 677.8 (1229.3) | 1886.9 (2044.6) |  |
| **Range** | 59.4 - 7333.764 | 25.8 - 7333.8 | 25.8 - 7333.8 | 67.5 - 7333.8 |  |
| **YLK 40** |  |  |  |  | < 0.001 |
| **Mean (SD)** | 11895.3 (25154.8) | 1212 (1799.3) | 9757.9 (26760.5) | 26643.3 (51209.5) |  |
| **Range** | 33.8 - 164096.8 | 15 - 12468.8 | 15.001 - 164096.8 | 15 - 164096.9 |  |
| **NGAL** |  |  |  |  | < 0.001 |
| **Mean (SD)** | 148.524 (213.3) | 56.472 (70) | 170.488 (284.4) | 478.661 (475.6) |  |
| **Range** | 3.2 - 1455.6 | 3.2 - 479.5 | 3.2 - 1455.6 | 8.3 - 1455.6 |  |
| **UMOD** |  |  |  |  | < 0.001 |
| **Mean (SD)** | 3300 (2638.9) | 3716.9 (2978.9) | 2494.4 (2465.7) | 3324.9(3310.2) |  |
| **Range** | 447.7 - 13611.9 | 250.3 - 13611.9 | 250.3 - 13611.9 | 250.3 - 13611.9 |  |

In terms of biomarker composites, individuals with AKI in subphenotype 1 were characterized by the highest mean troponin T levels recorded among all clusters. Additionally, biomarkers of cardiac congestion (pro-BNP) and remodeling (ST2) were also higher compared to subphenotype 2. Biomarkers of systemic (IL superfamily) and kidney inflammation such as TNFR1, TNFR2, were comparatively lower to subphenotypes 3 and 4. However, AKI patients in this subphenotype presented higher KIM-1 levels compared to subphenotype 2. AKI patients in subphenotype 2 were characterized by low levels of systemic and organ-specific biomarkers of inflammation and injury. Notably, these individuals had the highest levels of the anti-inflammatory biomarker, IL-13. AKI patients in subphenotype 3 were characterized by a marked influence of biomarkers of congestion compared to biomarkers of cardiac injury. Also, patients with AKI had predominantly low concentrations of biomarkers of systemic inflammation, but higher concentrations of TNFR1 and TNFR2 than subphenotypes 1 and 2. Interestingly, biomarkers of kidney injury such as IL-18, KIM-1 and NGAL were low and comparable to those levels exhibited by patients in subphenotype 2. Patients with AKI subphenotype 4 were the youngest and characterized by high concentrations of systemic (IL-1, IL-8, TNF-gamma) and kidney-related (TNFR1, TNFR2, YKL-40, MCP-1) inflammatory biomarkers. Similarly, they expressed higher levels of KIM-1 and NGAL levels when compared to subphenotype 2 and 3. They also expressed the lowest levels of cardiac injury activity. Full descriptions of biomarker composites per cluster are reflected in **Table 2** and visualization of selected clinical and biomarker signatures are seen in **Figure 4**.

**Figure 3.**
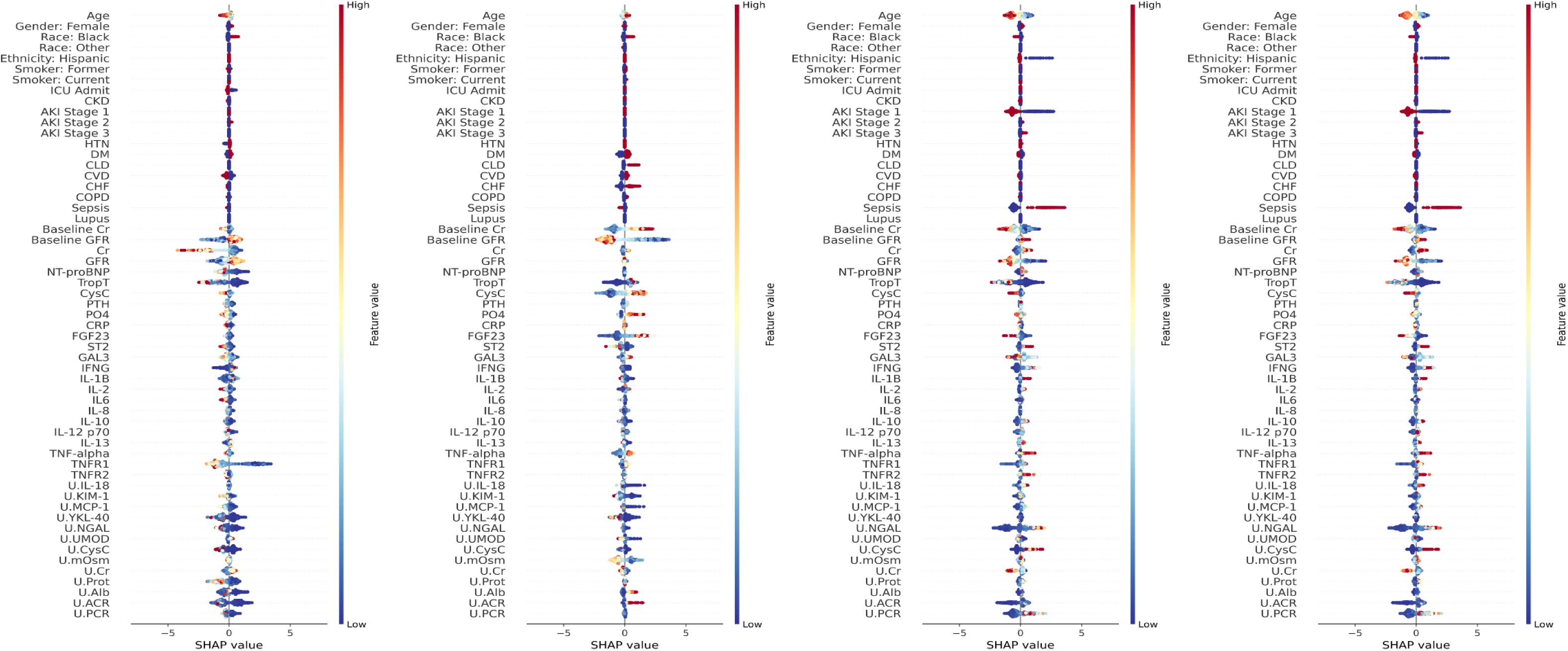
SHAP diagram comprising all variables included in subphenotype discovery and modeling.

**Figure 4.**
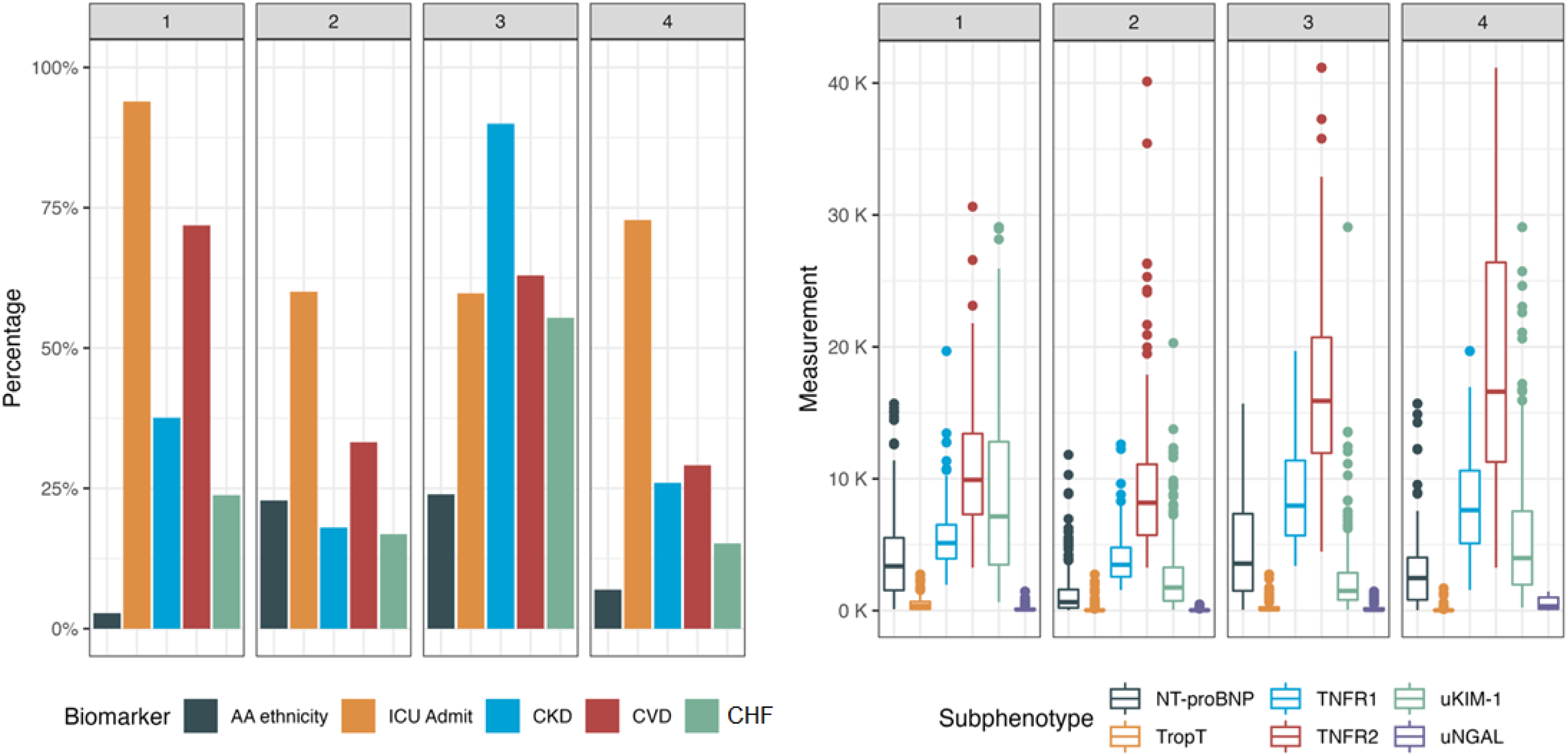
Key clinical characteristics (left) and selected biomarker composites (right) showing statistically significant differences between each subphenotype.

### Association between AKI subphenotypes and clinical outcomes

Over a median of 4.8 years of follow-up, there were 38 mortality events per 1000 person per year, 38.2 CKD events per 1000 person per year and 38.8 CVD events per 1000 person per year. Event rates (per 1000 person-year follow-up) per each subphenotype are shown in **Table 3**. AKI subphenotype 3 and 4 were independently associated with an aHR of death at 2.9 (95% CI: 1.8 – 4.6, p<0.001). In terms of CKD outcomes, subphenotype 3 was independently associated with an aHR of 2.6 (95% CI: 1.6 – 4.2, P<0.001) in fully adjusted models (**Table 3**). Furthermore, subphenotype 3 was independently associated with the risk of CVD events with an adjusted HR of 2.6 (95% CI: 1.6 – 4.1, P<0.002). **Figure 5** shows the forest plot of the subphenotypes and HRs for each outcome. **Supplemental figures 6-7** show different K-M curves for each outcome and subphenotype.

**Figure 5.**
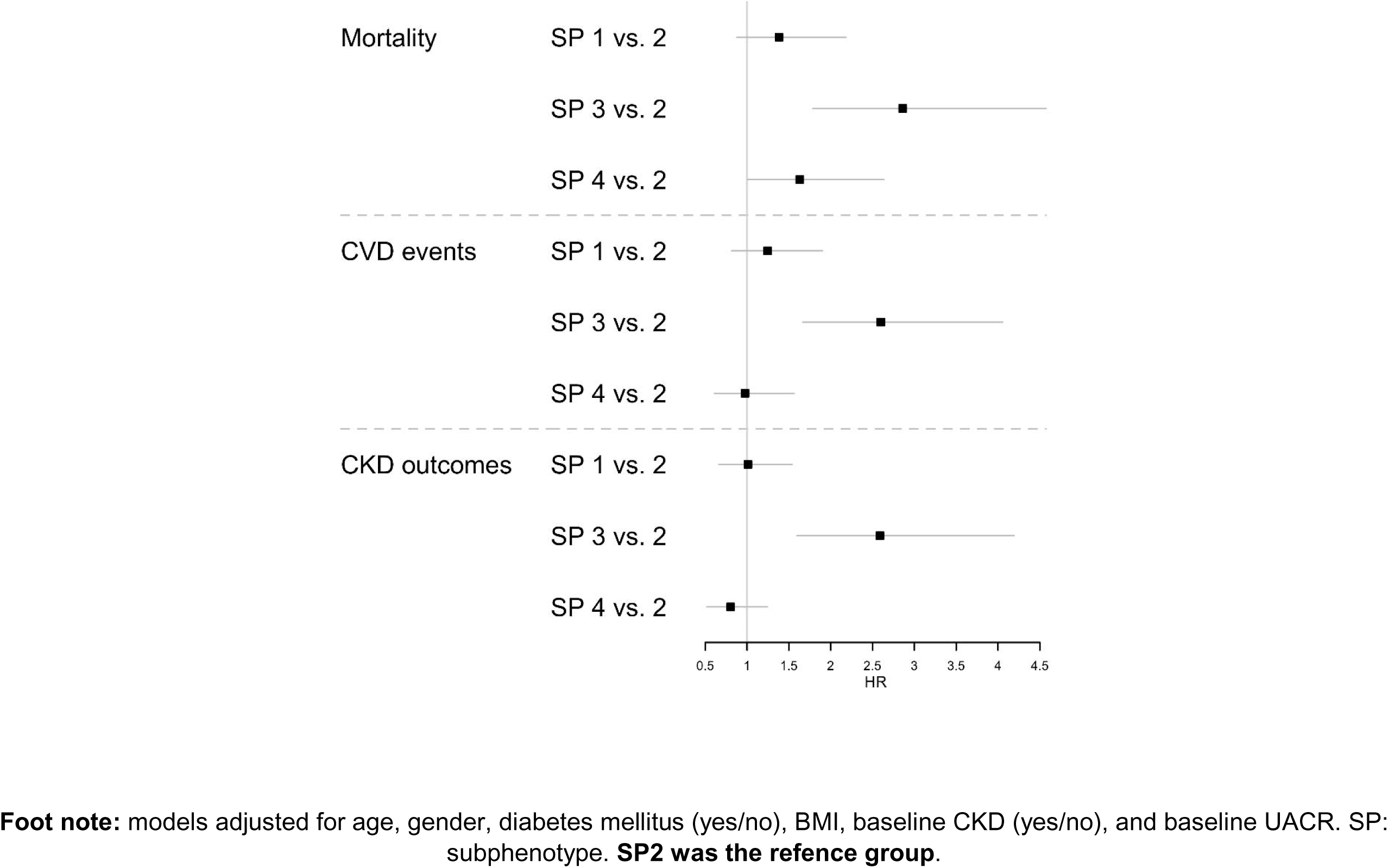
Forest plot analysis showing the 95% confidence interval adjusted hazard ratios for all outcomes among all subphenotypes.

**Table 3.** Hazard ratios (95% CI) for each subphenotype and the risks of longitudinal clinical events.

| Subphenotype | Event rate<br>per 1000<br>PY | Mortality |  |  | Event rate<br>per 1000<br>PY | CKD outcomes |  |  | Event rate<br>per 1000<br>PY | CVD events |  |  |
| --- | --- | --- | --- | --- | --- | --- | --- | --- | --- | --- | --- | --- |
|  |  | Adjusted models |  |  |  | Adjusted models |  |  |  | Adjusted models |  |  |
|  |  | HR | CI (95%) | p value |  | HR | CI (95%) | p value |  | HR | CI (95%) | p value |
| SP2 (ref) | 26.3 | 1 |  |  | 30.8 | 1 |  |  | 29.9 | 1 |  |  |
| SP1 | 39 | 1.4 | 0.8 - 2.2 | 0.16 | 33.3 | 1 | 0.7 - 1.5 | 0.9 | 41.9 | 1.2 | 0.8 - 1.9 | 0.3 |
| SP3 | 73 | 2.9 | 1.8 - 4.6 | <0.001 | 61.5 | 2.6 | 1.6 - 4.2 | <0.001 | 66.7 | 2.6 | 1.6 - 4.1 | <0.002 |
| SP4 | 29.4 | 1.6 | 1.01 - 2.6 | 0.04 | 37.5 | 0.8 | 0.5 - 1.2 | 0.3 | 28.4 | 0.9 | 0.6 - 1.6 | 0.9 |

**Foot note:** models adjusted for age, gender, diabetes mellitus (yes/no), BMI, baseline CKD (yes/no), and baseline UACR

## Discussion

We uncovered four clinically and molecularly distinct AKI subphenotypes through a series of unsupervised machine learning algorithms. These clusters are characterized by unique biological signatures as evaluated by a comprehensive panel of systemic and organ-specific serum and urine biomarkers reflective of different axes of disease and health. Biomarker composites were statistically different from each other and informed on the complexity of AKI syndromes through an unbiased approach. Furthermore, AKI subphenotype 3 and 4 were independently associated with 1.6 – 2.9-fold risk of death when compared with AKI subphenotype 2 (benign/reference cluster). Furthermore, AKI subphenotype 3 was independently associated with longitudinal CKD outcomes and CVD events in fully adjusted models among those patients who survived to hospitalization.

AKI has been defined on the basis of the KDIGO criteria using serum creatinine and urine output changes. While this tool has increased our capacity to identify AKI, there is vast evidence suggesting that molecular changes are present before meaningful serum creatinine and UOP changes are noticed clinically.^8, 36^ Furthermore, detectable tubular-interstitial abnormalities have been demonstrated to associate with AKI duration, dialysis needs and protracted kidney outcomes in large prospective studies.^37–41^ Moreover, the fine balance repair and maladaptive processes has been postulated to mediate the progression towards CKD.^42^ However, conceptualizing the complex nature of AKI and the intervening biological processes leading to distinct clinical trajectories is challenging for the clinician. This is in part due to large amounts of patient data but also due to the linear regressions used to evaluate the associations between individual or small set of biomarkers and outcomes. We demonstrated that machine learning algorithms allow us to deal with such data heterogeneity in an unbiased manner and discover clusters with unique clinical and molecular characteristics.

AKI subphenotype 1 (injury-predominant cluster) is characterized by senior patients with comparatively high levels of troponin T and KIM-1. Furthermore, these individuals presented higher levels of MCP-1 and YKL-40, which are markers of kidney and heart inflammation, compared to subphenotype 2 and 3. Yet, the majority of these patients had stage 1 AKI and had as low peak serum creatinine levels as compared to subphenotype 2 (benign AKI). These findings highlight the importance of incorporating additional tools for estimating kidney function in patients susceptible to muscle mass remodeling but also, to identify individuals at risk for clinical events.^43^ There are several clinical scenarios that could be represented by these findings, including patients with myocardial infarction or pulmonary embolism who are susceptible to hemodynamic instability, or patients with active cardiovascular disease undergoing contrast-based studies.

Patients in AKI subphenotype 3 had the highest prevalence of baseline cardiovascular disease, CKD, and heart failure. These patients presented the highest, pro-BNP, TNFR1 and TNFR2 levels and lowest baseline eGFR (38.9 ± 17 ml/min per 1.73 m^2^). Although these patients had the highest peak serum creatinine (3.4 ± 1.5) levels during hospitalization, they had low concentrations of urine IL-18, NGAL and KIM-1. A potential explanation relies in the time of collection of NGAL, which tends to have a short half-life compared to other biomarkers. Alternatively, we hypothesize that this scenario is plausible in the context of cardiorenal AKI, where creatinine fluctuations do not necessarily follow from acute injury marker elevations. AKI subphenotype 3 was independently associated with the worst outcomes such as death, CKD, and CVD events. TNFR1 and TNFR2 are relatively more stable markers of inflammation compared to other members of the TNF-alpha superfamily and elevated levels can be encountered in acute and chronic inflammation. Elevated TNFR levels has been found to be prognostic of cardiovascular disease, kidney disease progression and death among patients in the pre- and post- AKI stage as well as in patients with stable CKD with and without diabetes mellitus. As demonstrated in our study, patients these levels could be elevated and produced in the heart and vasculature while not necessarily driving worse AKI pathophysiology. Yet, given their capability to orchestrate diverse pathological cellular changes in the heart, kidney, vasculature, they have postulated as a redundant pathway leading to death. Patients in this cluster could resemble those with advanced heart and kidney disease, who present with cardiorenal syndrome. Conversely to prior data, it is possible that patients with creatinine fluctuations in the context of ischemic or hemodynamic changes (i.e. cardiorenal syndrome) without significant tubular injury activity could still present increased morbidity and risk of clinical events, mediated by these TNFR1 and TNFR2.

AKI subphenotype 4 characterized by patients with sepsis predominantly and AKI stage 3 (including dialysis dependency). Patients did not have a high prevalence of baseline comorbidities, but they did present the highest concentrations of biomarkers of systemic inflammation (IL-1, TNF-alpha, CRP), and markedly elevated levels of biomarkers of kidney inflammation (MCP-1, YKL-40) and injury (KIM-1, NGAL). This phenotype resembles individuals with septic shock who are at increased risk of complications such as death as reflected in fully adjusted models. While the burden of pre-existing comorbidities was not apparent in this group, the severe physiological stress elicited during a case of sepsis and the pathological pathways activated likely inform on a different pathophysiology and phenotype of AKI that requires more attention prospectively.

Our findings could potentially serve to gain insight on the diagnostic role and clinical relevance of biomarkers when incorporated into multi-dimensional models. Biomarker composites were statistically different within each subphenotype and denotes those different degrees of biological disturbances could identify the nature of AKI and inform on future clinical events. In the advent of new therapies that could potentially modulate inflammatory-predominant type kidney disease (i.e. SGLT2 inhibitors) or fibrosis (i.e. mineralocorticoid receptor antagonists), identifying these pathologic domains could be build up in process of exploring therapeutic targets in AKI or in the post-AKI stage; understanding that all AKI are not created equal. Our analyses were robust and validated through different unsupervised algorithms and our data builds upon previous analyses showing that AKI and CKD subphenotypes with can eventually lead to different outcomes. While these studies do not inform on underlying biological pathophysiology, our study is novel by incorporating biomarkers of key domains involved in kidney disease progression. Future studies may attempt to explore whether individuals within the same AKI subphenotype could share similar proteins and transcripts responsible of the biomarker composites and clinical outcomes evidenced in our cohort (multi-omic level). Using this clustering approach could assist with the challenges of stratifying AKI in different scenarios such as sepsis, acute cardiac injury, community-acquired AKI, and advanced organ disease.

By using a different approach (latent class analysis and K-means clustering), previous data has demonstrated the presence of two subphenotypes; one driven by congestive heart failure and benign biomarker profile and the other characterized by prevalent CKD and high concentrations of unfavorable biomarker profile when integrating 29 variables.^44^ The present study was characterized by more ample clinical and biomarker data integrating 53 variables, which could have resulted in different clustering representation. In detail, we applied several sophisticated methods to transform the raw data into different dimensionality, allowing us to use extensive mixed-type variables, and tackle two typical high dimensionality problems (sparseness and multiple chain imputations). These efforts turned into accessing granular subphenotypes where each subphenotype was well separated from the other, as shown in Figure 1 (dendrogram). In this context, it is possible that our 4 AKI subphenotypes ‘bisect’ the aforementioned 2 subphenotypes (congestion cluster with benign biomarker profile and CKD driven with unfavorable biomarker profile); thereby collapsing subphenotype 1 and 2 into one major cluster and subphenotype 3 and 4 into another single one. The angle of this study is unique and complementary to previous data, since we aimed to identify potentially useful ‘data patterns’ over statistically and numerally significant features that emulate real world data abundance. However, we acknowledge the potential trade with power reduction at the time of time to event analyses by deriving 4 subphenotypes instead of 2. The heterogeneity and clinical relevance found in the present study should be confirmed in additional studies with larger sample sizes.

In terms of limitations, due to the design of the ASSESS-AKI study, only patients who survived to the 3-month follow-up visit were included in the longitudinal data collection and analyses. Thus, there is some selection bias present. However, patients who die in-hospital or immediately following hospitalization are not in need of subphenotyping, and risk-stratifying for long-term outcomes. Our subphenotypes may not inform on those with the highest risk profile. Also, despite the heterogeneity of our AKI population (medical/surgical floors, ICU), a high proportion AKI cases were stage 1, which could carry a lower risk of CKD events compared to more severe stages. Most of our patients were white and we did not have a validation cohort, which limits the generalizability of our subphenotypes especially given the uniqueness of the biomarkers tested and the study design. Since this is data-driven approach, data pattern aggregation and cluster membership depend on the input of data. Therefore, if the input variables were to be modified, we could expect different AKI subphenotypes. Furthermore, reproducing these 4 novel clusters within each participating center in ASSESS-AKI is challenging, and potentially explained by the participating population in each center. For example, it may be difficult to reproduce AKI subphenotype 1 (cardiac/kidney injury predominant) in the surgical floor or AKI subphenotype 4 (injury-inflammation predominantly in the context of sepsis) in a cardiac care unit. Also, while our clusters describe altered biological pathways, they do not inform on the pathophysiology of AKI and genomic/transcriptomic analyses are needed. Moreover, cluster analysis summarizes patients’ heterogeneity of large data sets into discrete categories, which may result in loss of information in exchange for better clinical interpretation. Finally, subphenotype discovery depend largely on data input and feature characteristics, resulting in different feature aggregation and clinical/biomarker composites. Therefore, these subphenotypes may not necessarily produce in other settings (e.g. COVID-19) and further studies are needed.

We discovered 4 novel and clinically meaningful and statistically different AKI subphenotypes that inform on potential biological pathway abnormalities that associate with different risks for adverse clinical events and death. To the best of our knowledge, our data are novel as it comprehensively assessed the biological and statistical role of a comprehensive panel of biomarkers when obtained during the diagnostic phase of AKI. Biomarker composites within each cluster informed on the complexity of AKI syndromes through an unbiased approach. This study provides a novel look to the role of biomarkers in the diagnosis of AKI and their association to clinical events via multidimensional analyses.

## Supporting information

Strobe check list for cohort studies

## Data Availability

All data produced in the present study are available upon reasonable request to the authors

## Authors’ contributions

GVR* and WO* contributed equally to the elaboration and intellectual content. GVR, WO, SL, SGC and GNN participated in the design of the study and analysis of the data. GVR drafted the manuscript. All authors made substantial intellectual contributions to the manuscript and approved the last version of it. *Co-first authors.

## Acknowledgement

The authors would like to thank all of the ASSESS-AKI study participants, research coordinators, and support staff for making this study possible. The ASSESS-AKI was supported by cooperative agreements from the NIDDK (U01DK082223, U01DK082185, U01DK082192, and U01DK082183). We also acknowledge funding support from NIH grants R01HL085757, R01DK098233, R01DK101507, R01DK114014, K23DK100468, R03DK111881, R01DK093771, K01DK120783, P30DK079310, and P30DK114809. The ARID study was funded by a grant from Kidney Research UK (RP13/2015). The funder did not have any role in study design, data collection, analysis, reporting, or the decision to submit for publication.

## Supplemental Materials

**Supplemental Figure 1.**
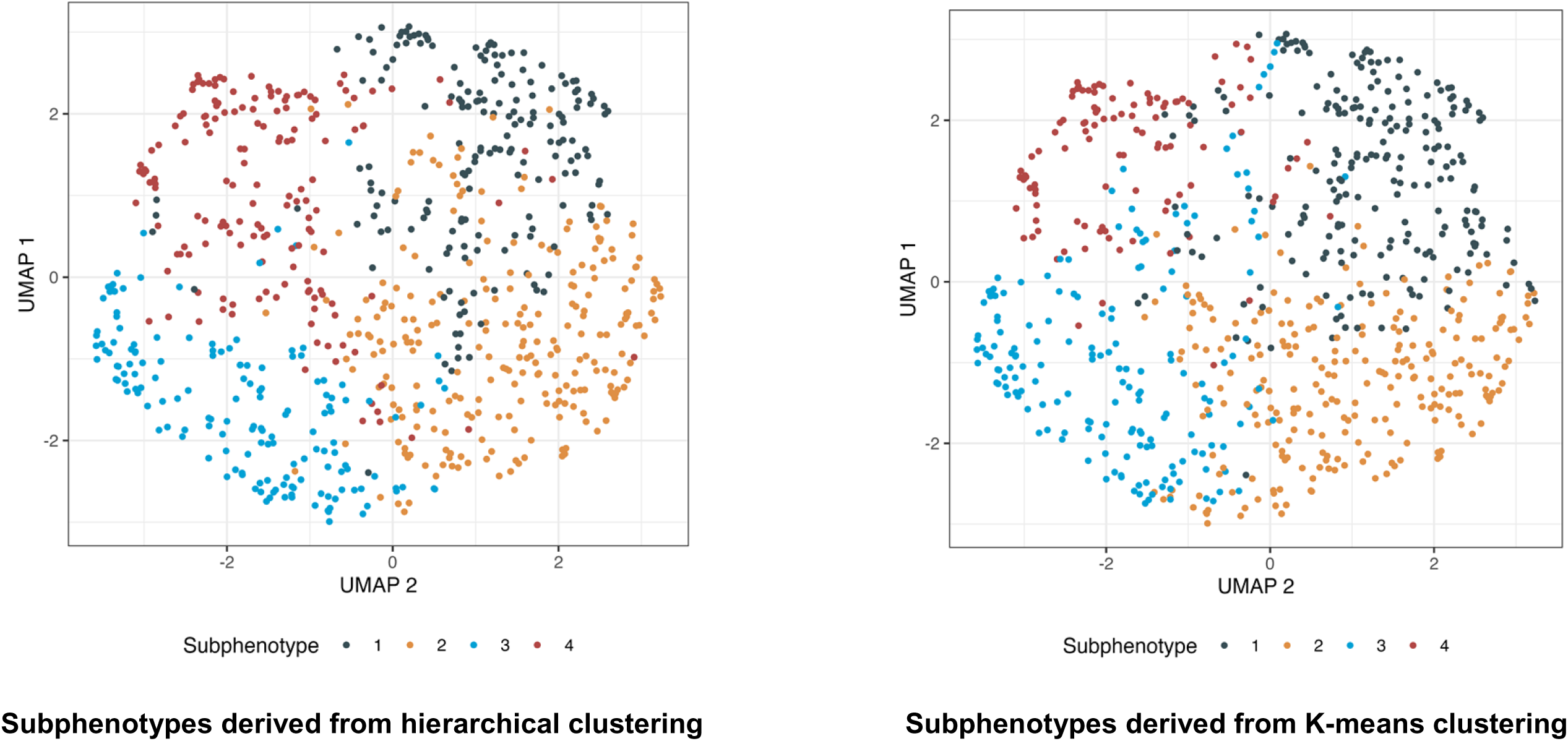
Hierarchical clustering (left) vs. K-means clustering (right) to demonstrate robustness of four subphenotypes.

**Supplemental Figure 2.**
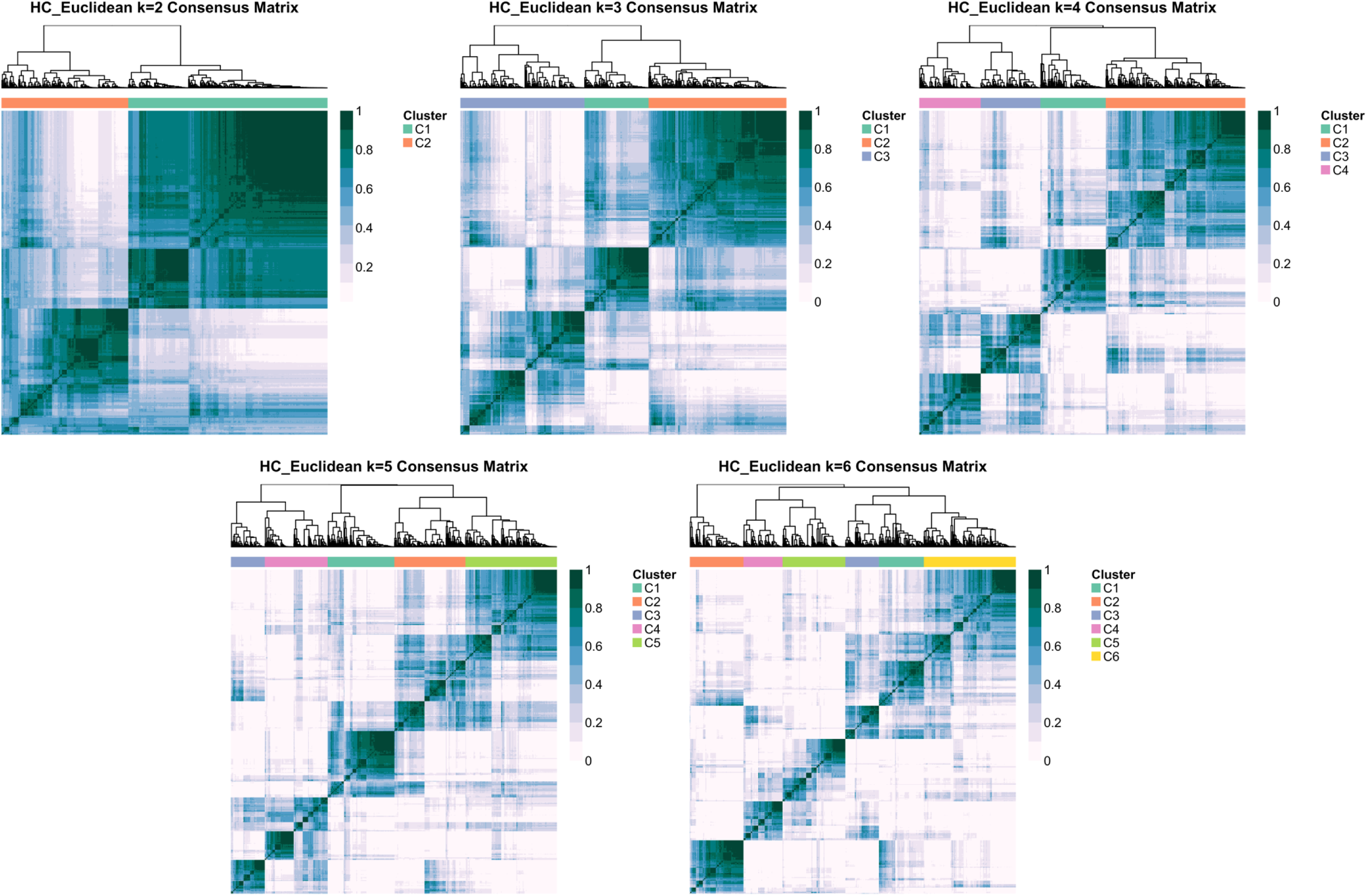
Consensus clustering demonstrating 2-6 subphenotypes.

**Supplemental Table 1.** Contingency table, consensus clustering (Y axis) versus hierarchical clustering (X axis)

| Single | 1 | 2 | 3 | 4 |
| --- | --- | --- | --- | --- |
| 1 | 136 | 45 | 0 | 0 |
| 2 | 8 | 241 | 1 | 0 |
| 3 | 0 | 28 | 131 | 0 |
| 4 | 6 | 6 | 5 | 141 |

**Foot note:** 45/181 (24.8%) patients in subphenotype 1 belong in subphenotype 2, from consensus clustering analyses. 28/159 (17.6%) patients in subphenotype 3 belong in subphenotype 2 via this approach. Individuals in AKI are clearly separated from other subphenotypes. Column: consensus clustering, Row: hierarchical clustering

**Supplemental Figure 3.**
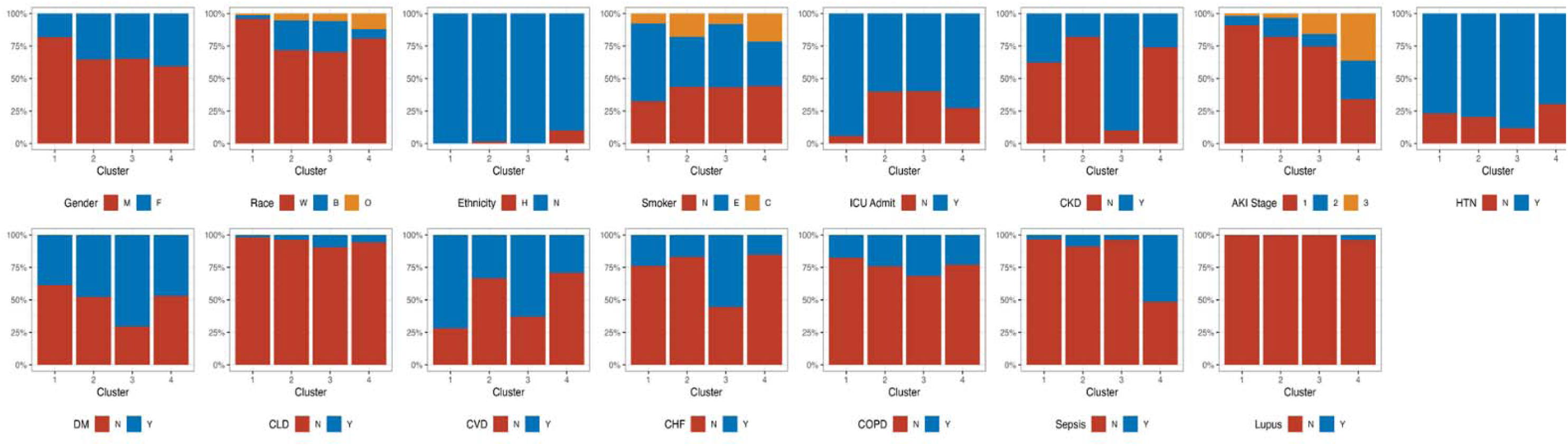
Cluster description based on categorical features.

**Supplemental Figure 4.**
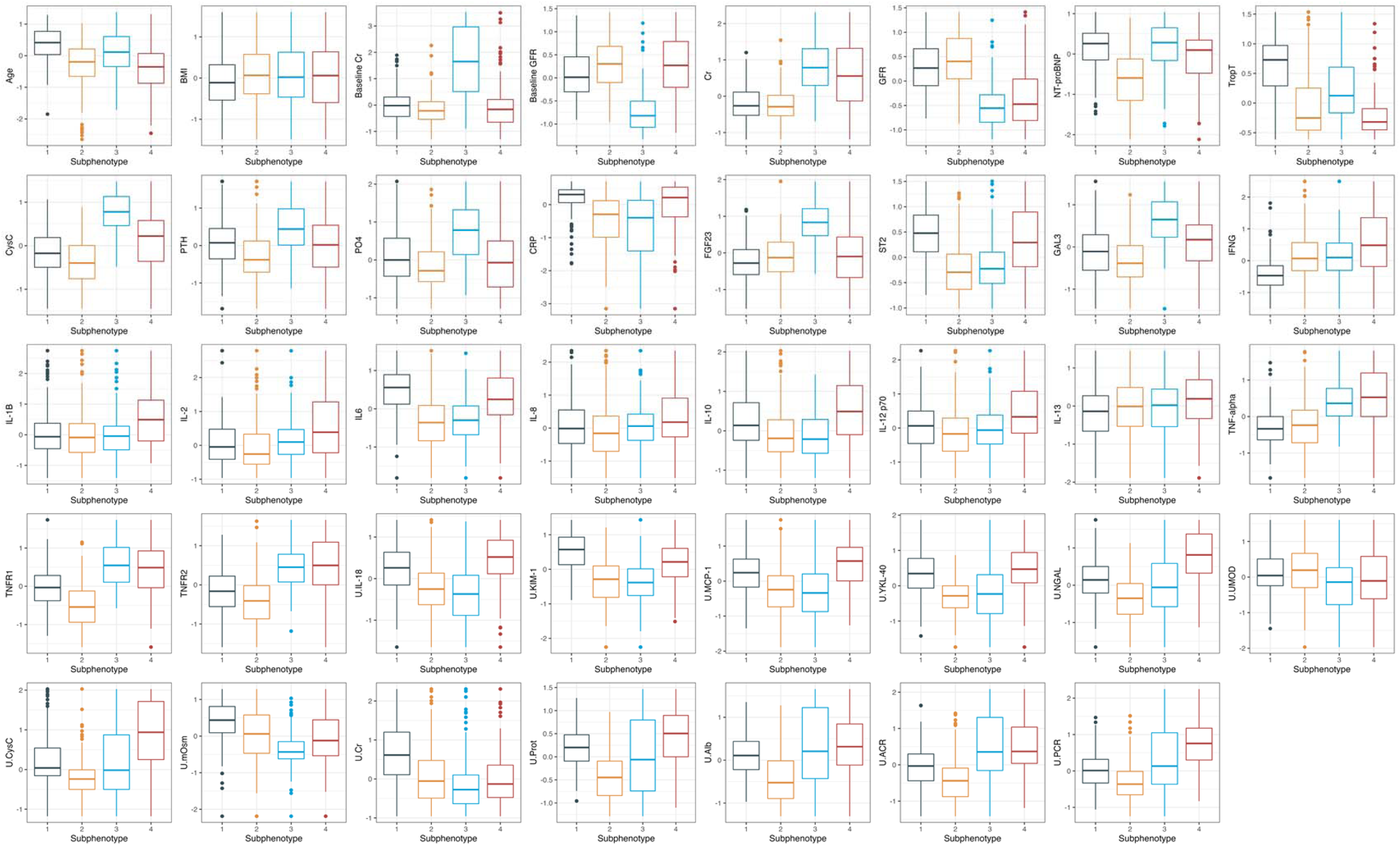
Cluster description based on numeric features with robust scaler transformation.

**Supplemental Figure 5.**
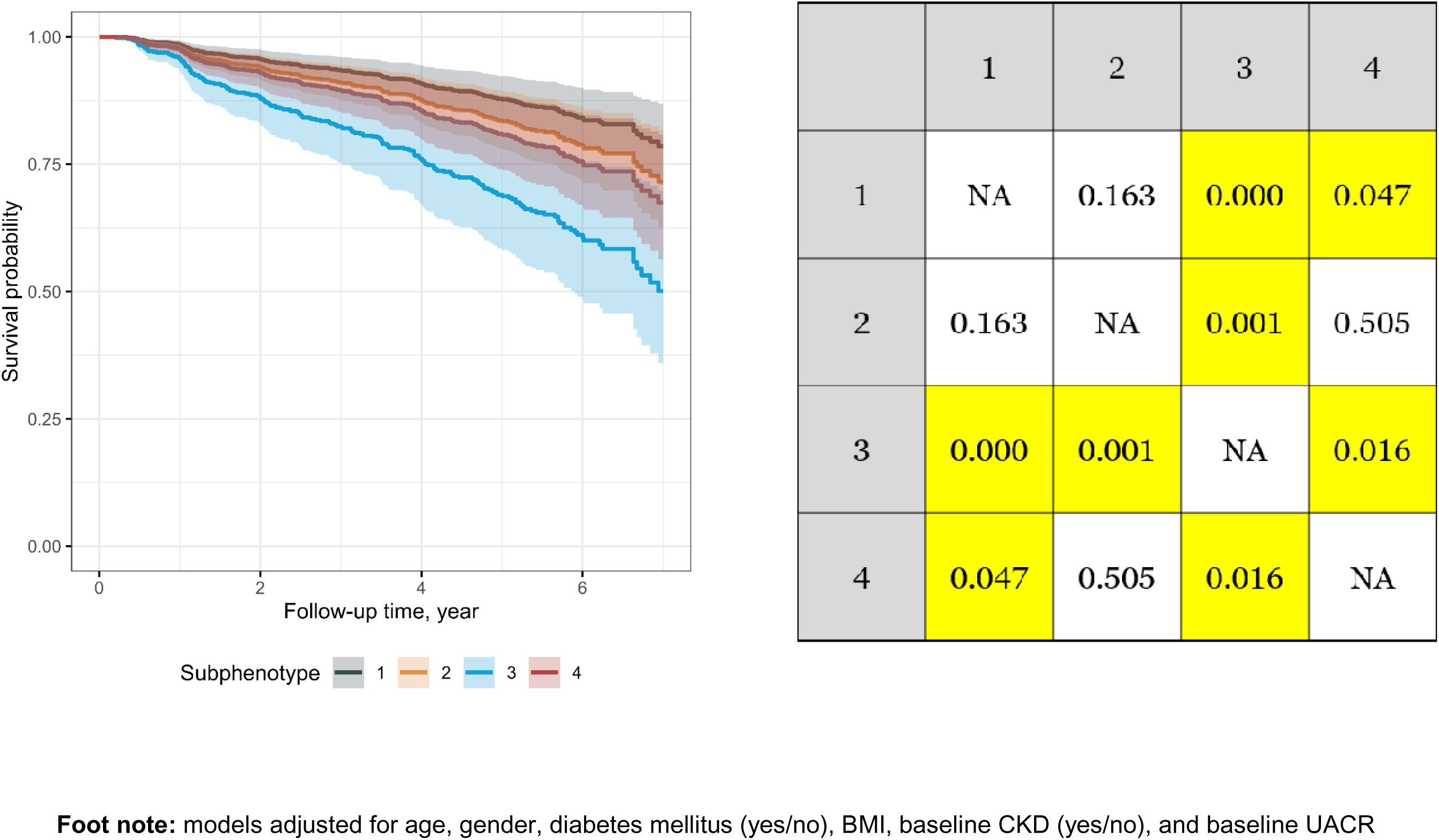
K-M and cumulative incidence curves for mortality per each subphenotype in adjusted models.

**Supplemental Figure 6.**
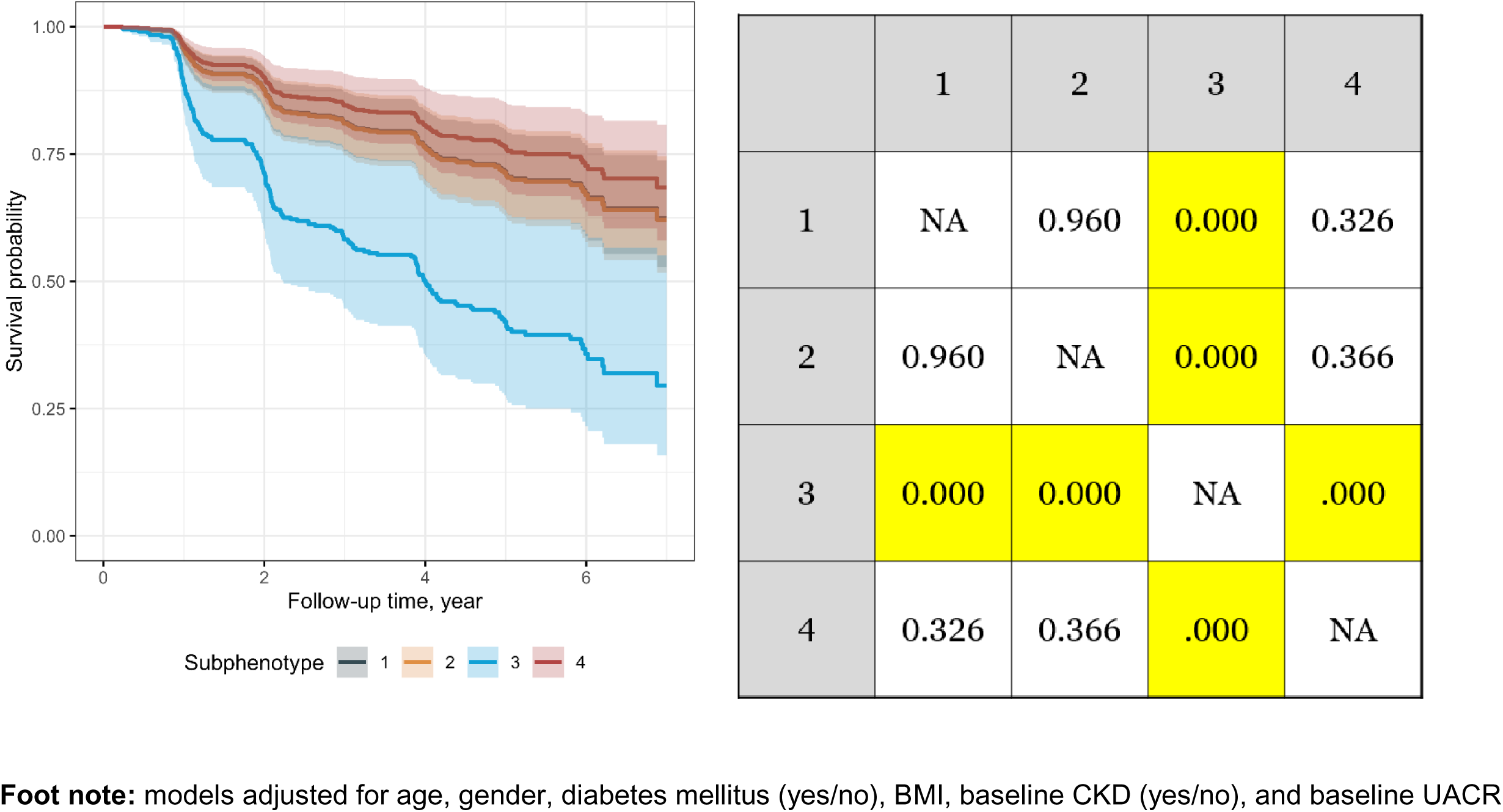
K-M and cumulative incidence curves for CKD outcomes per subphenotype in adjusted models.

**Supplemental Figure 7.**
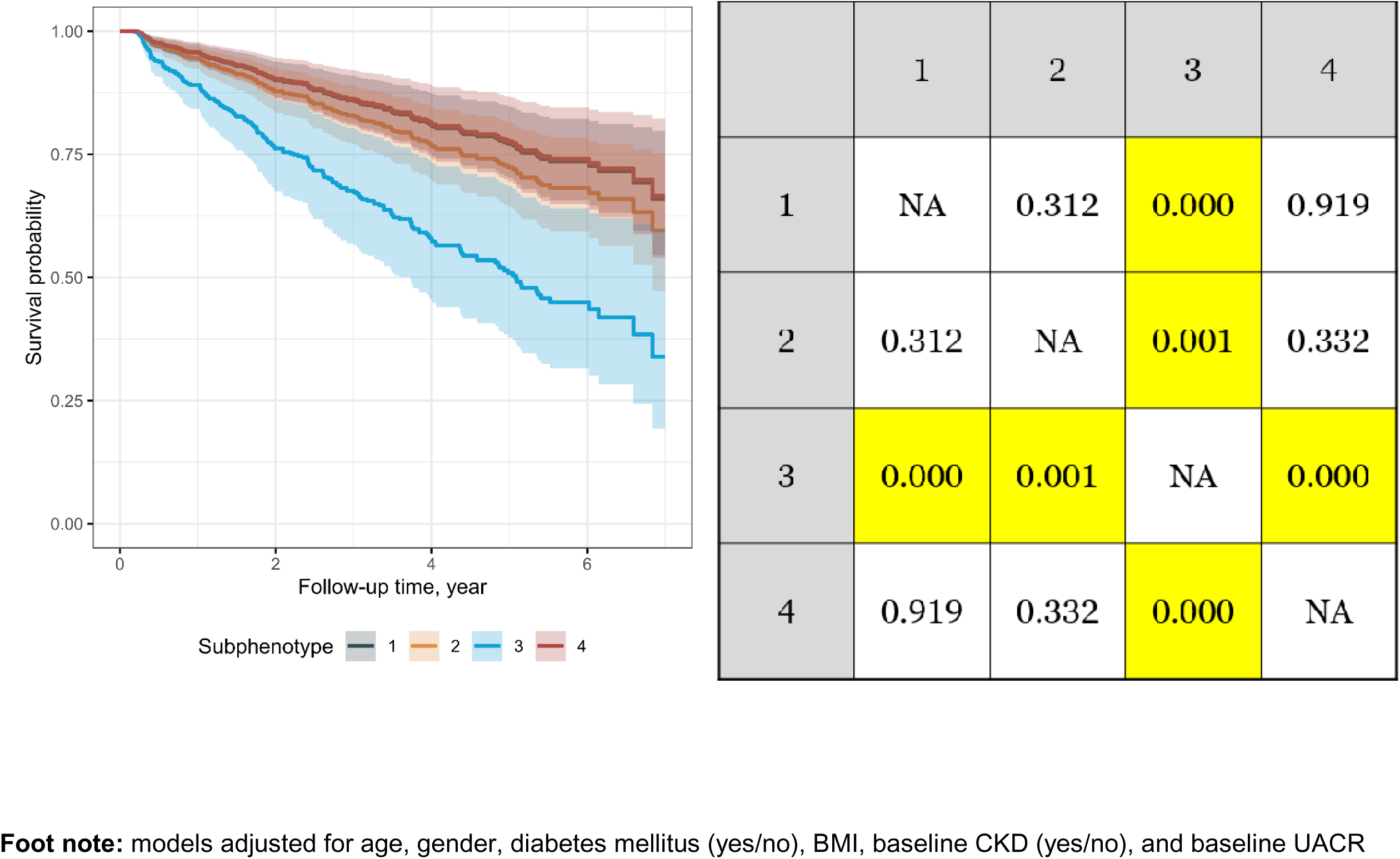
K-M and cumulative incidence curves for CVD events per each subphenotype in adjusted models.

